# Pre-analytical delay of blood cultures which hinders the management of sepsis and fosters the emergence of antimicrobial resistance is an adverse effect of laboratory centralisation

**DOI:** 10.1101/2025.06.09.25329298

**Authors:** Malila Noone

## Abstract

**Background:** The clinical diagnosis of sepsis is based on non-specific criteria and blood culture remains the gold standard confirmatory test. While early results are of wide clinical benefit delayed reports lead to prolonged anti-microbial therapy which fosters the emergence of antimicrobial resistance. Pre-analytical delay of a blood culture delays or decreases the chance of a positive report and a maximal four-hour delay between collection and incubation of the specimen is recommended by the United Kingdom Standards for Microbiology Investigations (UK SMI). This retrospective observational study documents compliance with this quality standard by National Health Service (NHS) hospitals across England. An appraisal is undertaken of the policies which governed laboratory centralisation and their impact on the microbiology service.

**Methods:** Freedom of Information (FOI) applications were submitted to 116 NHS Trusts in England requesting retrospective audit data showing compliance with the recommended pre-analytical standard for blood cultures. Information relating to the configuration of microbiology services and global laboratory costs were also requested.

**Results:** Reports were received from 89 Trusts (76.7%) comprising 146 acute hospitals. Only four hospitals (2.7%) showed full compliance with the four-hour pre-analytical standard. Service configurations varied widely. The anticipated savings resulting from centralisation have not been realised.

**Conclusions:** There was poor compliance with the quality standard for pre-analytical delay of blood cultures. Evidence is presented to show that the poor compliance rates reported are a result of the approach taken and the guiding policies applied when laboratory centralisation was imposed by NHS England. Reversal of these adverse effects will require mandatory implementation of UK SMIs and computing the cost of quality measures in the context of the overall health care benefit to the patient.

## Introduction

Sepsis is associated with high mortality and life changing after effects. This serious condition has been brought to public attention by Surviving Sepsis and other campaigns [1-3]. Reports of a rise in recorded sepsis deaths in the United Kingdom is a grave concern [4].

The clinical diagnosis of sepsis is based on reported symptoms, subjective clinical criteria and non-specific laboratory indicators. Blood culture remains the gold standard confirmatory test. When sepsis is suspected, early initiation of antimicrobial therapy can be lifesaving and the current practice of administering a cocktail of antimicrobial agents on an empirical basis covering the most likely causative pathogens has proved to be effective [5, 6]. A positive blood culture facilitates targeted antimicrobial therapy, aids accurate tailoring of adjunctive therapeutic agents and curtails investigations and clinical interventions which might otherwise have been undertaken. Identification of the specific infecting agent serves as a prognostic indicator and impacts on further clinical support measures as mortality risk and clinical progress varies across organisms,

The speed of the report is of critical importance. The sooner the laboratory identifies the specific pathogen, the sooner can treatment be de-escalated to the most effective narrow spectrum antimicrobial agent or to more effective agents if initial empirical therapy is found to be sub-optimal. The likelihood of the latter has increased with the growing prevalence of bacterial antimicrobial resistance [7, 8]. When early reports are expedited, there is a demonstrable impact on mortality rates and health care costs [9, 10]. While rapid reports are of wide clinical benefit, delayed reports lead to needlessly prolonged broad spectrum therapy which increases the likelihood of harmful side effects and fosters the emergence of antimicrobial resistance.

The current continuous-monitoring automated blood culture systems facilitate provisional reports within 08-18 hours of procuring a blood culture when collection and management of the specimen is optimised [11]. Blood cultures are time sensitive and widely used guidelines caution: “microbes grow, multiply, and die very quickly. If any of those events occur during the pre-analytical specimen management processes, the results of analysis will be compromised and interpretation could be misleading” [12]. The effect of delay on pathogens is variable and there is no ideal pre-analytical storage temperature. Room temperature and 37^0^C and are both inimical to pathogen survival and may lead to a negative report. If pathogens survive and grow during the delay, incubation may be followed by a lag in growth detection and a delayed positive reading or a false negative reading. Pre-analytical delay, as occurs when blood cultures are taken after the hours of operation of the laboratory, results in a reduced yield [13, 14]. On the other hand, reducing the pre-analytical interval increases detection rates and decreases time to detection [15, 16]. Minimising delay also enhances the validity of a negative result which will permit withdrawal of antimicrobial agents after clinical review. A negative result may be discounted and lead to prolonged antimicrobial therapy with its associated iatrogenic harms unless the validity of a negative result is assured by demonstrating minimal pre-analytical delay.

The pre-analytical delay standard recommended in the United Kingdom Standards for Microbiology Investigations (UK SMI) S12 is that inoculated bottles should be loaded onto an automated blood culture analyser without delay and ideally within a maximum of 4 hours from time of collection [17]. Compliance with this standard serves as a measure of the quality of the microbiology service and its ability to support the diagnosis and management of sepsis.

This retrospective observational study documents the degree of compliance by hospitals in England with the recommended pre-analytical standard for blood cultures. An appraisal is undertaken of the policies guiding laboratory centralisation in England and their impact on clinical microbiology standards. Proposals for reversing the adverse effects of laboratory centralisation are presented.

## Materials and Methods

### Setting and Context

The NHS is a publicly funded healthcare system in the United Kingdom (UK) comprising NHS England, NHS Scotland and NHS Wales. NHS England (NHSE) provides healthcare to a population of over 57 million in England. In 1990, acute care hospitals in England were set up as independent, administrative units termed ‘Trusts’. Since then, many of the original single hospital Trusts have merged to form larger administrative units, still termed Trusts, often incorporating several hospitals and sometimes establishing inter-Trust networks.

When pathology services in England were reviewed in 2006, international comparisons showed that the UK pathology service was one of the best value systems in the world with expenditure per capita on in-vitro diagnostics about half that of equivalent countries in Europe and a quarter of the expenditure on diagnostics in the USA. The use of the alternative term ‘’Laboratory Medicine’’ was advocated and it was suggested that pathology should be managed as a core clinical service in its own right, taking the cost of an individual test in the context of overall health care costs and the benefit to the patient [18]. Nevertheless, in 2008, after a further review, “back office and pathology services” (categorised as being comparable) were ear-marked for speedy centralisation [19]. Centralisation was driven by the potential for savings with a target reduction of global pathology (all clinical laboratory specialities) expenditure from approximately 4% to <1.6% of total Trust expenditure. The favoured configuration was a ‘hub and spoke’ model which concentrated non-urgent work at a centralised hub to achieve benefits through economies of scale. Multidisciplinary “blood science” laboratories were to operate on-site in each acute hospital throughout a 24-hour period to achieve a four-hour turnaround time (TAT) for urgent blood specimens while microbiology laboratories were often moved off-site.

### Study Design

Freedom of Information (FOI) requests were submitted to 116 acute-care non-specialist NHS Trusts in England. The submission was addressed to the designated FOI officers in each Trust with requested information available from the laboratory management team. The principal question was the percentage of blood culture sets incubated within four hours of specimen procurement at each acute hospital in the financial year 2022/23 [Fig 1]. In June 2022 an NHS England directive to all Trusts referred to the UK SMI S12 and reinforced the requirement for prompt attention to blood cultures [20]. This implied that recent audit results were readily available. It was assumed that all Trusts were using automated blood culture analyser systems.

Additional questions helped clarify the configuration of the microbiology service. The standard opening hours of each laboratory was not documented. The global pathology (all clinical laboratory specialities) cost as a percentage of total Trust annual expenditure for the same year was also requested. This is a standard measure which is included in Trust annual financial reports.

The UK Freedom of Information Act 2000 provides right of access to a wide range of information held by public authorities, including the NHS. Information disclosed under the FOI Act is protected by copyright. Copyright protected material was not presented in this review. A request made, as a courtesy, for anonymised publication of data with safeguards against identification of the individual Trust was refused by one Trust.

Application for ethical approval was not considered necessary as patient data was not requested.

No funding was received by the author.

## Results

Of the 116 administrative units (Trusts) receiving the FOI request, 15 Trusts did not respond and gave no reason for withholding information. (Response rate 87%). Although Trusts are legally obliged to respond to FOI requests, failure to answer was not pursued in consideration of the heavy clinical workload faced by many Trusts. A further 12 Trusts stated that they were unable to provide compliance data as their pathology service was in the hands of an off-site provider. An appeal was made to several FOI officers that the request should be brought to the attention of the designated Trust clinical microbiologist responsible for the pre-analytical handling of specimens, since an off-site provider would only be responsible for the analytical service. This did not change the stand taken.

Compliance data was received from 89 Trusts. (Response rate 76.7). A request made for anonymised publication of data was refused by one Trust. Results are therefore reported for 88 Trusts.

### Service Configurations

The responses received and information available in the public domain were used to construct service configurations and arrangements for processing blood cultures. (Table 1). Results are presented only for acute hospitals which are defined as those providing 24 hour acute-care services and submitting more than 500 blood cultures annually. Fifty nine Trusts designated ‘independent Trusts’ did not transport blood cultures off site and did not appear to receive cultures from other Trusts. Thirty independent Trusts running a total of 35 acute hospitals had traditional arrangements for blood cultures with on-site facilities for incubation and processing of specimens in each hospital. Hub and spoke arrangements were established in 29 independent Trusts running a total of 69 acute hospitals. There were 12 inter-Trust networks incorporating 25 Trusts running a total of 35 acute hospitals with hub and spoke arrangements. Of 63 spoke hospitals, 39 transported specimens directly to the central hub while 24 had satellite automated blood culture analyser units for incubation of cultures. Only bottles flagging positive were transported to the central hub laboratory to be processed. In one of these spoke hospitals, specimens flagging positive within 18 hours were tested using a BioFire Blood Culture Identification Panel before transfer of the specimen to the hub laboratory. Spoke to hub distances for the 63 spoke hospitals ranged from 1.9-40miles (median 10miles). Four Trusts used commercial off-site service providers.

**Table 1:** Trusts and hospitals: service configurations.

|  | Trusts | Acute Hospitals | Hub or on-site incubation & culture | Spoke: satellite unit on site. Positives to hub | Spoke: transport direct to hub |
| --- | --- | --- | --- | --- | --- |
| Independent Trusts<br>1-2 hospitals each | 30 | 35 | 35 |  |  |
| Independent Trusts<br>2-6 hospitals each<br>Hub & Spoke | 29 | 69 | 32 | 11 | 26 |
| Inter-Trust networks<br>Hub & Spoke | 25 | 35 | 15 | 9 | 11 |
| Off-site commercial<br>provider | 4 | 6 |  | 4 | 2 |
| Total | 88 | 145 | 82 | 24 | 39 |

### Audit of compliance with Pre-Analytical Delay Standards

Thirty five Trusts (39.7%) had no compliance data for 2022/23. Fifty three Trusts reported compliance data but there was no consistency in compliance rates between hospitals run by a single trust or between hospitals in an inter-trust network. Compliance rates are therefore presented for each of the 145 hospitals in 88Trusts (Table 2).

**Table 2:** Compliance with pre-analytical standards in 145 hospitals.

| Category | 100% | 80-99% | 50-79% | <50% | No data | Total |
| --- | --- | --- | --- | --- | --- | --- |
| Hubs or on site culture | 3 | 24 | 25 | 9 | 21 | 83 |
| Spokes with satellite unit | 1 | 12 | 3 | 4 | 4 | 24 |
| Spokes –transport only |  | 6 | 4 | 19 | 10 | 39 |
| **Total** | **4** | **42** | **32** | **32** | **35** | **145** |

Since the request did not ask for laboratory opening hours, it was not possible to determine which hospitals were able to incubate blood cultures throughout a 24-hour period. Full compliance was reported by only four hospitals (2.7%). Three of these were central hubs with facilities for on-site incubation and culture and one was a spoke hospital with facilities for incubation in a satellite automated analyser. A compliance rate of 80-99% was reported by 42 hospitals (28.9%) and is suggestive of incubation of cultures beyond routine laboratory hours. Only 14 of 88 Trusts (15.9%) reported undertaking regular audit of pre-analytical delay. Of the 35 Trusts unable to provide compliance data, 12 were planning regular audits and expecting to comply with the recommended standards in the future - two by installing satellite units. Reasons given for the lack of data included lack of access to or inability to interrogate the computer, and inability to change computer settings to include time of specimen procurement. Only one trust reported inclusion of an explicit comment in the final report indicating that the blood culture was received in the laboratory more than four hours after it was taken and that the delay may reduce the chance of growing relevant pathogens.

All trusts followed the current UK SMI 12 except for two Trusts which followed UKSMI B37 which preceded it. Compliance with the four-hour standard was denoted as only “aspirational” in the latter and one Trust reported that compliance audit was not undertaken for that reason.

### Laboratory Service Cost As a Percentage of Trust Cost

Costs were provided by 70 Trusts (79.5%). Eighteen Trusts withheld cost data citing it commercially sensitive or confidential. Financial arrangements between Trusts and with outsourced providers were variously described as a “Joint Venture Partnership”, “Integrated Pathology Partnership” or “Managed Pathology Service”. Overall, the reported interquartile range of costs was 2.95-4.13 (Median 3.3%). Hub and spoke arrangements did not appear to confer a cost advantage over independent Trusts with on-site incubation and culture showing median costs of 3.3 and 3.2 respectively. (Table 3)

**Table 3:** Costs by Trust Configuration.

| Category | Trusts | Cost reports | Costs withheld | IQ Range/<br>Median |
| --- | --- | --- | --- | --- |
| Independent Trusts<br>On- site incubation and culture | 30 | 27 | 3 | 2.95-4.0/ 3.2 |
| Independent trusts<br>Hub & Spoke configuration | 29 | 23 | 6 | 3.0-4.6/3.3 |
| 3. Inter-Trust networks<br>Hub & Spoke configuration. | 25 | 17 | 8 | 3.0-4.7/3.7 |
| 4. off-site commercial | 4 | 3 | 1 | Not available |
| Total | 88 | 70 | 18 | 2.95-4.13/3.3 |

## Discussion and Conclusions

Compliance with the four hour pre-analytical delay standard for blood cultures as recommended in UK SMI S12 is an auditable measure of the ability of a microbiology service to deal with blood cultures in a timely manner thereby optimising support for the diagnosis and management of sepsis. Of a total of 146 acute hospitals managed by 88 administrative units/Trusts in England, only four hospitals (2.7%) reported full compliance with this standard.

Poor compliance with this quality measure is a predictable result [21] of the policies which governed laboratory centralisation in England. The variation in compliance rates between hospitals and the diverse service configurations clearly demonstrate the absence of a cohesive plan for microbiology services in particular.

The chief quality measure proclaimed as a benefit of laboratory centralisation was a reduction in analytical turn-around times (TAT) which related mainly to specimens processed in on-site multidisciplinary “blood science” laboratories. This directive failed to recognise that although the standard analytical TAT for final microbiology reports ranged from three to five days, the specimens themselves required timely handling with robust internal and external transportation systems and access to laboratory facilities throughout the day. Although standard opening hours of each laboratory was not documented in this study only four appear to provide 24-hour access. A further 42 (28.9%) showed >80% compliance suggestive of partial access after routine opening hours. Restricted access inevitably leads to pre-analytical delay of blood cultures. It has been suggested that the delay in handling blood cultures taken over the weekend when facilities were less than optimal may be linked to the increased mortality over the weekend branded the ‘’weekend effect’’ [14, 22].

Quality standards for microbiology are embodied in the UKSMIs and regular audit of procedural standards is recognised as good laboratory practice by the Royal College of Pathologists (RCPath) which has published surveys of blood culture practices in its Bulletins [23]. Although UK SMIs were incorporated into laboratory manuals they were not declared mandatory by NHSE and its standards were therefore not enforceable. As a result, auditable standards for microbiology were not widely incorporated into IT systems despite the investment in IT based connectivity which accompanied laboratory centralisation. Of 88 Trusts responding to the FOI application for compliance rates, 35 (39.7%) were unable to provide data for the previous year 2022/23, and only 14 (15.9%) reported undertaking regular compliance audits. Difficulties in interrogating the system and the inability to incorporate time of specimen procurement into the system indicates a lack of flexibility which is likely to arise when pathology or IT support is centralised or outsourced with long term 5-7 year contracts.

It should be noted that although UK SMIs have not been declared mandatory, once they are incorporated into a laboratory procedure manual, non-compliance may be legally challenged as is likely with increasing public interest in sepsis. Drawing attention to pre-analytical delay in the final report may offer some legal protection in this regard. Only one Trust included an interpretive comment referring to pre-analytical delay and its effect on the reliability of the result. Responsibility for maintaining and auditing quality standards clearly lies with the designated hospital clinical microbiologist and this may have ethical implications [24]. Referral of the FOI request to an off-site provider of analytical services by 12 trusts hints at a lack of direct control, a lack of ownership, disengagement with quality issues or a sense of alienation felt when the analytical laboratory service is moved off-site. **T**he lack of mandatory status for UK SMIs is likely to have further disempowered clinical microbiologists and curtailed their ability to secure the funding required in order to comply with quality standards especially in the face of prevailing cost controls.

Laboratory **c**entralisation was driven by the demand for cost savings but the potential contribution of the microbiology laboratory to the savings agenda was not critically scrutinised. Economies of scale can be demonstrated in areas benefiting from automation such as serological tests for specific infectious disease markers as in blood donor screening. These specimens are more robust, training procedures less complex, and the NHS is well placed to take advantage of discounts associated with bulk purchasing, single system contracts and leasing agreements. Economies of scale do not however readily translate to microbiological specimens requiring labour intensive “culture and sensitivity’’ tests. Centralisation of acute care microbiology was instituted despite it imparting little or no benefit in terms of direct financial savings. The imposed financial target of a reduction of global laboratory expenditure to <1.6% of total Trust expenditure and does not appear to have been realised. Laboratory costs reported by 70 Trusts ranged from 2.95-4.13/3.3% (median 3.3%) and the favoured hub and spoke arrangements do not appear to confer a cost advantage over single hospital sites with traditional arrangements which showed lower median costs of 3.2%. The substantial expenditure on approved specimen packaging, the open ended costs of specimen transport, and the relatively expensive “blood science” laboratories serving each hospital site are likely to have reduced the anticipated savings.

The adverse effects of centralising microbiology laboratories on test quality and its indirect impact on antimicrobial stewardship and hospital acquired infections and associated costs is addressed in a comprehensive report [25]. There have been no measures in place in England to monitor the impact of laboratory centralisation on quality aspects of healthcare such as duration of stay, costs, antibiotic use, emergence of antimicrobial resistance and healthcare associated infections. Consequently, the emergence of antimicrobial resistant organisms and other iatrogenic patient harms may not be recognised as attributable to laboratory shortcomings. Warnings about the widespread general overuse of antimicrobials masks the issue of the emergence of antimicrobial resistance in individual patients due to delayed reports leading to the prolonged use of broad spectrum antimicrobials.

Compliance with pre-analytical delay standards may be achieved by installation of satellite automated analysers if they are located at a site with 24-hour technical cover supported by adequate internal transport systems. Current advances in technology enables loading by non-specialist staff although this flexibility has to be negotiated and space constraints and training implications overcome. This however would be the first step in the pathway to providing speedy results. If positive bottles are left unread overnight or over the weekend or if bottles are not processed promptly in a 24-hour facility, delays in time to Gram stain and time to antimicrobial test results will continue to cause patient harm.

The use of molecular tests on positive bottles prior to further processing (as undertaken in one laboratory) would mitigate the effects of delay but only be in relation to the most common pathogens. Such procedures also provide rapid pathogen identification and detection of common resistance genes directly from EDTA blood samples which would speed up the diagnosis of sepsis. They are likely to be clinically cost effective but cannot entirely replace standard diagnostic microbiology which remains a “science of interpretive judgement that is becoming more complex’’ [12] and which is critical for the detection of rare and emerging pathogens and unusual antimicrobial resistance patterns. These new diagnostic systems do not support further centralisation as they require effective on-site clinical liaison which is more difficult when the service is centralised.

Microbiology should be recognised as an essential 24-hour service with facilities on-site for handling critical specimens such as blood cultures as well as other specimens from relevant sites. The latter may identify the source of sepsis and may be the only source of a positive isolate from a patient with sepsis. UK SMIs should have mandatory status and the cost of implementing their quality standards should take account of indirect savings and take “the cost of an individual test in the context of overall health care costs and the benefit to the patient” [18]. An adequate response to the Surviving Sepsis campaign requires a seamless and speedy pathway from specimen collection to the final report. Few recommendations have such a clearly proven impact on patient survival and on indirect financial benefit. It is also of considerable community benefit in terms of containing the emergence of antimicrobial resistance which is a worldwide concern.

## Data Availability

The UK Freedom of Information Act 2000 provides right of access to a wide range of information held by public authorities, including the NHS. The data presented are available from each NHS Trust in England by means of Freedom of Information requests. All data received are summarised within the manuscript.

## Limitations of the study

**1)** Responses to the Freedom of Information (FOI) request were accepted as factually correct and could not be confirmed by other means.

ii) The study was limited to England but is applicable in other countries where centralisation of laboratories has taken place or is being encouraged.

iii) Results relate to the period 2022/23. The institutions referred to in the study may have introduced improvements.

## Data availability

The data presented is available from each NHS Trust in England by means of FOI requests.

## Ethics Approval

Application not required as patient data was not used.

---

This is a FOI request relating to the microbiology laboratory service and blood cultures.

The data should be easily available by interrogating laboratory information systems or from audit reports.

I should be grateful for a response in the form detailed below.

The data will not be used in a way which will identify an individual Trust.

Thank you for your time.

Dr Malila Noone. Retired Microbiologist. No commercial interest

**<u>FREEDOM OF INFORMATION REQUEST: BLOOD CULTURES</u>**

1. Which Trust hospitals currently submit blood for culture?

i) Are any blood cultures incubated on site or sent directly to BCPS to be incubated and processed?

ii) If incubated or processed off site, what is the distance in miles between the site and the offsite laboratory?

iii) Number of bed days for all patients per acute site. Bed days= Finished Consultant episodes x average length of stay

2. Does the Trust follow the UK Standards for Microbiology Investigations(SMI S12 &) in relation to Blood Cultures? YES/NO

3. Do you undertake formal compliance audits of the recommended pre-analytical standard for blood cultures of a four hour TAT from collection to incubation? YES/NO

4. What percentage of blood culture sets achieved this standard in the year 2022/23 at each site.

If full compliance with the pre analytical standard cannot be met, are clinicians made aware of the risk via a statement in the report on specimens which encountered delay?

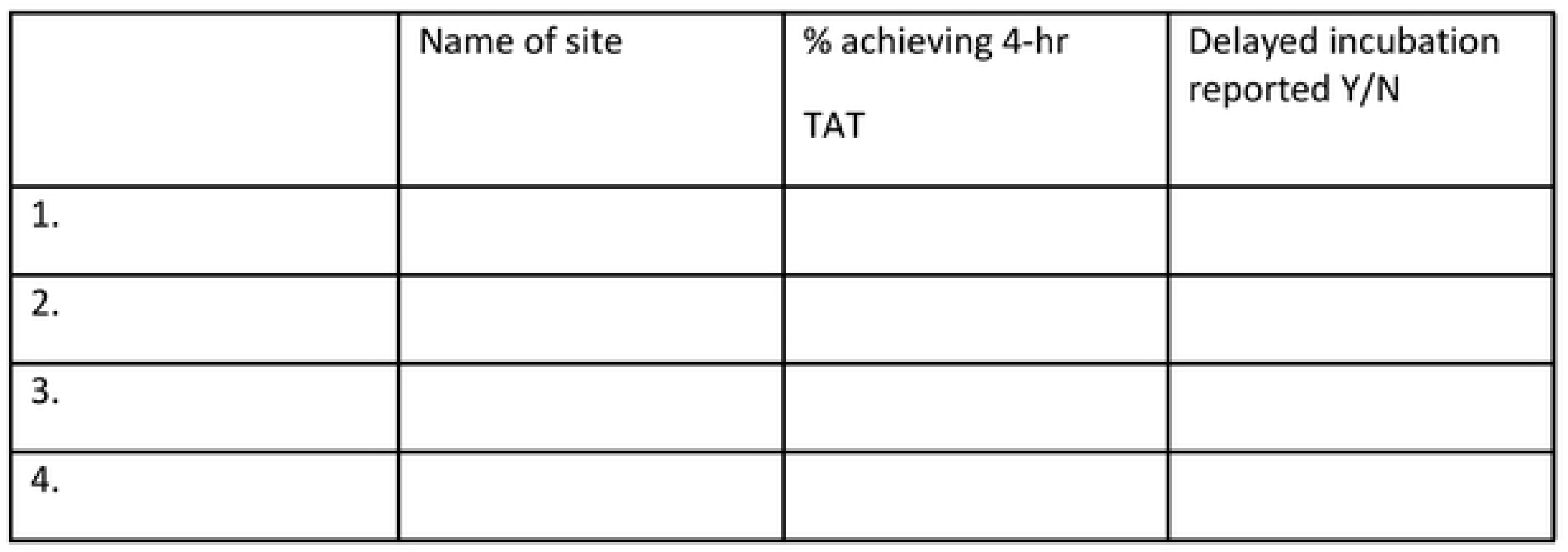

|  | Name of site | % achieving 4-hr TAT | Delayed incubation reported Y/N |
| --- | --- | --- | --- |
| 1. |  |  |  |
| 2. |  |  |  |
| 3. |  |  |  |
| 4. |  |  |  |

Comments:

5. What is the population served by the Trust?

6. What was the total expenditure on laboratory services (directorate of pathology) as a percentage of total Trust expenditure in the year 2022/23?

7. Please append a copy of your blood culture protocol or policy or other document which informs clinical staff when they should take a blood culture.

